# Non-replication of structural brain changes from Mindfulness-Based Stress Reduction: Two combined randomized controlled trials

**DOI:** 10.1101/2021.06.14.21258762

**Authors:** Tammi RA Kral, Kaley Davis, Cole Korponay, Matthew J. Hirshberg, Rachel Hoel, Lawrence Y Tello, Robin I Goldman, Melissa A Rosenkranz, Antoine Lutz, Richard J Davidson

## Abstract

Studies purporting to show changes in brain structure following the popular, eight-week Mindfulness-Based Stress Reduction (MBSR) course are widely referenced despite major methodological limitations. Here, we present findings from a large, combined dataset of two, three-arm randomized controlled trials with active and waitlist (WL) control groups. Meditation-naive participants (n=218) completed structural MRI scans during two visits: baseline and post-intervention period. After baseline, participants were randomly assigned to WL (n=70), an 8-week MBSR program (n=75), or a validated, matched active control (n=73). We assessed changes in gray matter volume, gray matter density, and cortical thickness. In the largest and most rigorously controlled study to date, we failed to replicate prior findings and found no evidence that MBSR produced neuroplastic changes compared to either control group, at either the whole-brain level or in regions of interest drawn from prior MBSR studies.

## Introduction

Research on mindfulness-based interventions has increased in response to a growing interest in alternative treatments for reducing stress and improving well-being. Yet, a recent meta-analysis found that the proportion of high quality publications in this domain have not improved over time, although there are a growing number of high quality studies being conducted (Goldberg *et al*., 2018). Moreover, findings from a few small studies have permeated popular media with the notion that a few weeks of training in Mindfulness-Based Stress Reduction (MBSR) can lead to measurable changes in brain structure (Congleton, Hölzel and Lazar, 2015; Schulte, no date), despite a lack of replication or confirmatory analysis of these findings in a fully randomized trial, and reliance on a single measure of structural neuroplasticity.

MBSR is a popular, manualized mindfulness intervention that was originally developed for use in clinical settings to improve patients’ ability to cope with pain (Kabat-Zinn, 1982; Kabat-Zinn, Lipworth and Burney, 1985). MBSR is efficacious for ameliorating symptoms of multiple psychopathologies (Wielgosz *et al*., 2019) and for reducing stress (Chiesa and Serretti, 2009). Studies have begun elucidating cognitive and neural mechanisms underlying mindfulness training-related changes in affect (Desbordes *et al*., 2012; Hölzel *et al*., 2013; Kral *et al*., 2018), cognition (Jha *et al*., 2010; Chiesa, Calati and Serretti, 2011; Mrazek *et al*., 2013; Gallant, 2016), and pain (Gard *et al*., 2012; Zeidan and Vago, 2016), among other processes. Studies have also examined whether mindfulness meditation practice leads to changes in brain structure, as described in a meta-analysis by Fox et al (2014), in light of numerous studies demonstrating changes in brain structure following behavioral training in other domains (Draganski *et al*., 2004, 2006; Colcombe *et al*., 2006; Ilg *et al*., 2008). However, only three studies included in the meta-analysis assessed changes specifically following training with MBSR. The majority of included studies focused on cross-sectional research of long-term meditation practitioners from a variety of meditation traditions, who may have pre-existing differences relative to non-meditators, and idiosyncratic lifestyle factors associated with engaging in long duration meditation practice and meditation retreats. Conversely, MBSR is a standardized, manualized intervention in which participants receive similar training over an 8-week period, where pre-post design research can control for individual differences at baseline. Thus, we focus the current investigation specifically on the effects of MBSR. Prior research has reported that participants who completed MBSR had increased gray matter density (GMD) in the hippocampus, posterior cingulate cortex (PCC), temporo-parietal junction (TPJ), cerebellum, and brainstem (Hölzel *et al*., 2011), and increased gray matter volume (GMV) in left caudate (Farb, Segal and Anderson, 2013). While prior research on MBSR lacked measures of cortical thickness (CT), research on meditation more broadly has reported regional increases in CT, including in the insula (Fox *et al*., 2014). More recent studies of the impact of short-term mindfulness meditation training on brain structure have consisted of pilot trials, with fewer than 15 subjects and no control group (e.g., Yang *et al*., 2019).

However, prior studies on MBSR-related changes in brain structure had marked limitations. These included a lack of active control groups and randomization, reliance on circular analysis (Kriegeskorte *et al*., 2009), and small sample sizes – methodological limitations that are prevalent in meditation research more broadly (Chiesa and Serretti, 2009; Davidson and Kaszniak, 2015). The current study aimed to address these limitations by integrating a waitlist and a well-matched active control group with larger sample sizes (i.e., a minimum of 70 participants per group), in a set of two, rigorous, randomized controlled trials with a pre-post designs from which we created a combined dataset. In this way, we were able to test for structural changes that were specific to mindfulness meditation training, rather than non-specific effects associated with wellbeing interventions more generally. Recent literature also stresses the need for replication (Ioannidis, 2005; Moonesinghe, Khoury and Janssens, 2007; Button *et al*., 2013), especially for meditation research (Fox *et al*., 2014).

Prior research used varied measures of structural neuroplasticity, including GMV, GMD and/ or CT. GMV provides a measure of the size of a region of interest (ROI) in mm^3^, whereas GMD indicates the concentration of gray matter within an ROI (or within each voxel). CT indicates the thickness of the cortical sheet between the white matter and pial surfaces, and thus is not available for sub-cortical regions. All three measures can be estimated with voxel-based morphometry using information about the deformation of voxels between native and group space. Surface-based analysis can also be used to calculate GMV and CT using information derived from geometric models of the cortical surface. Growth in any of these measures putatively reflects the same underlying processes, including synaptogenesis and gliogenesis (Zatorre, Fields and Johansen-Berg, 2012). However, some prior research has found that surface-based analysis was more effective (e.g. with higher sensitivity and lower variability) for subcortical volume estimation (i.e., in error-prone regions) (Dewey *et al*., 2010), provided the best estimates for change over time in longitudinal models (Clarkson *et al*., 2011), and had better sensitivity to detect differences in brain volumes between clinical groups compared to a tensor-based analysis pipeline (Schmitter *et al*., 2015). Therefore, we used a surface-based analysis pipeline for the present study, though we subsequently re-processed the data using tensor-based methods as well, for sensitivity analysis (see Supplementary Information).

Changes in brain structure in association with mindfulness meditation training would provide evidence of structural neuroplasticity and may elucidate potential mechanisms underlying benefits of mindfulness meditation. In prior work, hippocampus and insula were selected as ROIs *a priori*, due to their role in emotion control and awareness (respectively), their activation during meditative states, and prior associations with long-term meditation training and increased GMV in these regions (Hölzel *et al*., 2011). The insula, amygdala and anterior cingulate contribute to the salience network, which is associated with emotional reactivity and subjective awareness processes that are hypothesized to change with mindfulness training (Seeley *et al*., 2007; Lutz *et al*., 2015). PCC and TPJ are major nodes within the default mode network, which is implicated in self-referential thought and mind-wandering (Spreng, Mar and Kim, 2008; Fox *et al*., 2015), that have been shown to change with mindfulness training (Jain *et al*., 2007; Mrazek *et al*., 2013). Given the evidence for mindfulness-related changes in function and psychological processes associated with these brain regions (Brewer *et al*., 2011; Allen *et al*., 2012; Desbordes *et al*., 2012; Creswell *et al*., 2016; Fox *et al*., 2016; Kral *et al*., 2018; Wielgosz *et al*., 2019), structural changes in gray matter might also be expected. Indeed, prior research provides evidence for mindfulness-related changes in brain structure in the default mode and salience networks, among other regions (Hölzel *et al*., 2011; Farb, Segal and Anderson, 2013; Fox *et al*., 2014).

We attempted to replicate prior findings of increased GMD following MBSR in hippocampus, PCC, TPJ, cerebellum, and brainstem, and increased GMV in caudate. In addition to the aforementioned regions, we also assessed structural changes in the amygdala and insula, as these regions are involved in affective processing that may change with mindfulness training (Allen *et al*., 2012; Desbordes *et al*., 2012). While prior research did not find associations between structural changes and amount of MBSR practice time, we tested for such associations, given the broader range of MBSR practice time in the current study relative to prior work. We hypothesized that MBSR practice time would be associated with increased GMV, GMD and CT in all ROIs except amygdala, where we expected an inverse relationship between size and practice time, given the inverse relationship between MBSR-related reductions in amygdala GMD and stress in prior research (Hölzel *et al*., 2010). Thus, the current research sought to replicate and extend the literature on structural neuroplasticity associated with short-term mindfulness meditation practice in MBSR.

## Results

### Whole-brain analysis

There were no significant group differences for change in brain structure (GMV, GMD, or CT) for MBSR compared to the Health Enhancement Program (HEP) active control group, or the waitlist (WL) control group, in the whole-brain analysis. This is consistent with a prior whole-brain analysis of GMV conducted with sample one (Korponay *et al*., 2019). There were no significant interactions between MBSR and HEP practice time and change in GMV in the whole-brain analysis. Un-thresholded statistical maps are available on NeuroVault (Gorgolewski *et al*., 2015) at this link: https://indentifiers.org/neurovault.collection:7634.

### ROI analysis

There were no significant group differences for change in brain structure for MBSR compared to HEP or WL for any of the ROIs (*p*s>0.1). The non-significant result for right amygdala is depicted in Figure 1a. See Supplementary Table 1 for results of statistical tests of change in GMV for all ROIs. Results are consistent regardless of the inclusion or exclusion of influential outliers. Non-significant results of sensitivity analysis using multiple imputation to account for missing data, as well as analysis of GMD from SPM12 and SPM-CAT12 are presented in Supplementary Tables 3-8.

**Figure 1.**
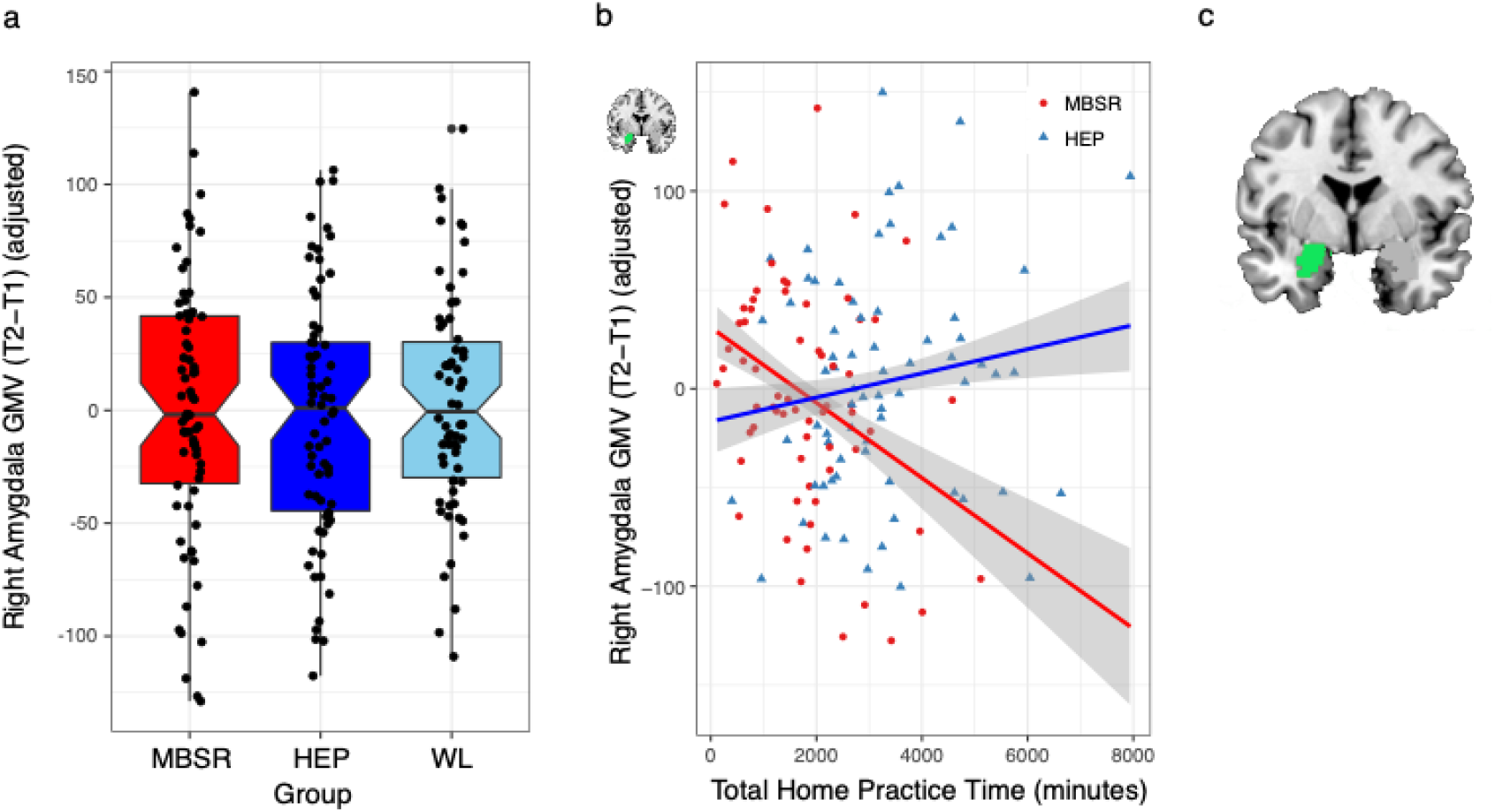
Right amygdala GMV change and MBSR practice time. **(a)** There were no significant differences between groups in right amygdala gray matter volume (GMV) change. **(b)** MBSR practice was related to reduced right amygdala GMV significantly more than HEP practice. **(c)** FreeSurfer anatomical label from aseg for right amygdala (green). Error envelopes represent 1 standard error above and below the point estimates of the means, the dependent variables are adjusted for covariates (e.g., age, gender, sample, and total brain GMV), and adjusted data points are overlaid.

### MBSR practice time

MBSR participants practiced at home an average of 32 hours (standard deviation=20 hours, range=2–85 hours), and HEP participants practiced at home an average of 56 hours (standard deviation=33 hours, range=7–255 hours). MBSR participants attended 8.14 of 9 possible classes, on average (range = 4 to 9 classes), and HEP participants attended 8.44 of 9 possible classes, on average (range = 2 to 9 classes). Significant effects of MBSR practice time were limited to the amygdala, and relationships with the other 8 ROIs were non-significant (*ps*>0.05). MBSR practice time was associated with reduction in right amygdala volume significantly more than HEP practice (*t*(128)=-3.30, *p*=0.001, *p\**=0.01, p*η*^2^=0.08; Figure 1b). See Supplementary Table 2 for results of all statistical tests examining the impact of practice time on change in GMV. However, the group by practice time interaction was trend-level or non-significant in the sensitivity analyses using multiple imputation and GMD (see Supplementary Tables 4, 6, and 8).

### Mindfulness

We examined self-reported mindfulness based on the FFMQ to gauge the effectiveness of the MBSR intervention. A prior study reported on the results for sample one, whereby MBSR was associated with increased mindfulness (*p*<0.05, within-group) that marginally differed from WL (*p*=0.09), but did not differ from HEP (*p*=0.33) (Goldberg *et al*., 2015). When collapsing across both studies, results were consistent with the prior report, whereby MBSR differed significantly from WL (*t*(208)=-2.70, *p*=0.01), but not from HEP (*t*(208)=-1.01, *p*=0.31), p*η*^2^=0.05. Across both samples, mindfulness increased following MBSR (*t*(70)=3.86, *p*<0.001, p*η*^2^=0.18) and HEP (*t*(65)=3.39, *p*=0.001, p*η*^2^=0.15).

## Discussion

The current study sought to replicate and extend the functional significance of prior work demonstrating increased GMD following mindfulness meditation training in hippocampus, posterior cingulate, cerebellum, brainstem, and temporoparietal junction (Hölzel *et al*., 2011), and increased GMV in caudate (Farb, Segal and Anderson, 2013). We combined two datasets to yield sample sizes of 70 or more participants per group. Both datasets were collected with the same rigorous methods and three-arm randomized controlled trial design, using MBSR, a well-matched, active control (HEP), and a waitlist control. We expected to replicate prior results of increased GMD following short-term MBSR training in hippocampus, caudate, TPJ, and PCC, and reduced volume for amygdala, and we also hypothesized that these effects would be larger for participants who spent more time practicing mindfulness meditation. We failed to find any group differences in GMV, GMD, or CT in support of these hypotheses.

It is unlikely that the failure to replicate prior work was due to ineffective training. The MBSR intervention was effective with regard to expected changes in neural, psychological, and cognitive outcomes: MBSR reduced amygdala reactivity and increased amygdala-VMPFC functional connectivity to emotional stimuli in sample one (Kral *et al*., 2018), increased PCC resting functional connectivity with dorsolateral prefrontal cortex in sample two (Kral *et al*., 2019), and increased self-reported mindfulness (reported in Goldberg et al., 2015 for sample one, in addition to the results presented here). The active control intervention, HEP, also increased self-reported mindfulness, and MBSR participants did not differ significantly from HEP on this measure. The current study lends evidence that MBSR-related improvements in self-reported mindfulness may not be specific to mindfulness meditation practice, but rather, related to other aspects of the course that are common to similar interventions (e.g., benefits from learning well-being skills from experts in HEP; for an in-depth discussion see Rosenkranz et al.) (Rosenkranz, Dunne and Davidson, 2019). Participants in the current study were at least as engaged with the MBSR coursework as in the prior research, if not more engaged, based on the time they reported practicing meditation at home (which ranged from 2 to 85 hours with a mean of 32 hours in the current study, compared to a range of 7 to 42 hours and a mean of 23 hours in prior work) (Hölzel *et al*., 2010).

Despite the lack of group differences in change in regional brain structure, we observed a significant interaction of group (MBSR versus HEP) and practice time on change in right amygdala GMV. The more time participants spent practicing MBSR outside of class, the larger their reduction in right amygdala volume following the intervention compared to practice with HEP, the active control intervention. On average, participants with less than 27 hours of total MBSR practice time had no change in amygdala volume, and the lower bound of the confidence interval at this point was 20 hours of total MBSR practice time (or an average of about 22 minutes per day). Therefore, practicing mindfulness meditation for less than 22 minutes per day for a few months is unlikely to lead to structural change in the amygdala. In addition, changes in the early stages of mindfulness meditation training, such as during MBSR training in previously untrained individuals, may be different than in later stages or for longer interventions. Along these lines, a recent, well-powered study assessing meditation training similar to mindfulness meditation, but with a 50% longer duration than MBSR, found significant increases in CT relative to two active control interventions in prefrontal cortex extending to anterior cingulate, and in bilateral occipital cortex extending to inferior temporal cortex (Valk *et al*., 2017). While the current study thus provides initial evidence that MBSR-related reductions in amygdala volume may depend on the degree of engagement with practice, the effect was small and failed to survive the sensitivity analyses. Thus, it should be interpreted with caution and warrants attempts to replicate in future work.

The results of the current study failed to support the hypothesis that short-term training in mindfulness meditation is associated with significant group differences in change in regional brain structure compared to a well-matched, active control intervention or a waitlist control group in an adequately powered, rigorous RCT design. Despite previous research suggesting that short-term mindfulness meditation training impacts the structure of the brain, results of the present study failed to replicate these group differences. While this highlights the importance of replication studies, it also raises new questions. There were important differences between the current study and prior work, including the populations from which participants were drawn and differences in the study design and methods. Prior work recruited participants who elected to participate in an MBSR course (Hölzel *et al*., 2011; Farb, Segal and Anderson, 2013), and were thus not randomly assigned, while the current study utilized a rigorous randomized controlled trial design. The participants in prior studies may have had more “room for improvement”, since they sought out a course for stress reduction, with some samples recruited specifically based on the presence of high stress in participants the month prior to study participation (Hölzel *et al*., 2010). In contrast, the current set of RCTs employed a relatively long list of inclusion/ exclusion criteria, including exclusion for use of psychotropic medication or psychiatric diagnosis in the past year, resulting in unusually healthy samples with, e.g., very low levels (or absence) of baseline anxiety and negative affect. While the RCT design employed here provides the strongest scientific methodology, this increased rigor likely comes at the expense of ecological validity – the simple act of choosing to enroll in MBSR may be associated with increased benefit.

It is notable that the current study also had sample sizes over 3 times that of prior work (e.g., n=75 MBSR participants in our final sample compared to n=20 or less participants per group in prior work) (Hölzel *et al*., 2011; Farb, Segal and Anderson, 2013). Given the low sample sizes of prior work, and the larger samples and lack of replication in the current study, there is a possibility that prior results suffered from inflated effect sizes and low positive predictive value (Button *et al*., 2013). For example, we found medium effect sizes (ranging from *Cohen’s d* = 0.39 to *d* = 0.43) for the significant group differences between MBSR and WL in self-reported mindfulness (from pre- to post-intervention), whereas the prior research found large effect sizes (ranging from *Cohen’s d* = 0.77 to *d* = 1.48). Moreover, the very small magnitude of standard effect size estimates for the non-significant group differences in change in regional GMV in the current study can be interpreted as “no effect” (e.g., partial *η*^2^ = 0.01 for the difference between MBSR and HEP for change in left hippocampus GMV). These null effects, in conjunction with our large sample size and rigorous matched comparison condition, allow us to conclude that MBSR had no effect on altering brain structure.

As more research is conducted on this topic, the importance of reporting results of replication attempts should be emphasized in light of known publication bias for positive findings (Ioannidis *et al*., 2014; Coronado-Montoya *et al*., 2016). Likewise, it is important for future research to examine individual differences in engagement and efficacy of MBSR, as well as the optimum length and duration of daily practice for a mindfulness meditation intervention to confer benefits. The lack of significant group differences between MBSR and control groups in the current study suggests that interventions lasting longer than the standard 8-week MBSR course might be required to confer changes in brain structure.

## Materials and Methods

The present study was registered with ClinicalTrials.gov, and combined data across two clinical trials (NCT01057368 and NCT02157766), which started approximately 5 years apart. Baseline data collection (T1) for sample 1 occurred between February 2010 and May 2011, and data collection following the intervention period (T2) occurred between June 2010 and October 2011. For sample 2, baseline data (T1) collection occurred between November 2014 and March 2017, and data collection following the intervention period (T2) occurred between March 2015 and July 2017. The experimental design was comparable across both data sets (see Supplementary Figures 1 and 2 for the CONSORT diagrams from each trial). Both clinical trials ended upon completion.

### Participants

Data were combined for meditation-naïve participants (MNP) from samples one (n=124, average age 48.1 ± 10.7 years, 79 female) and two (n=139, average age 44.1 ± 12.7 years, 82 female). Sample size for each data set was determined with a power analysis. We recruited healthy human participants within the Madison, WI community using flyers, online advertisements, and advertisements in local media for a study of “health and well-being” or the “benefits of health wellness classes.” Participants were included if they were adults between 18 and 65 years old with no prior training or formal practice in meditation or mind-body techniques (e.g., Tai-Chi), or expertise in physical activity, music, or nutrition. Participants were excluded from enrollment if they had used psychotropic or nervous system altering medication, a current diagnosis of sleep disorder, a psychiatric diagnosis in the past year, any history of bipolar or schizophrenic disorders, history of brain damage or seizures, or a medical condition that would affect the participants ability to safely participate in study procedures. There were no differences in socio-economic status between MBSR and either control group based on the Hollingshead Index (*p*s>0.10) (Hollingshead, 1975).

Following baseline data collection, participants were randomized to one of three groups using a stratified block assignment procedure to ensure age- and gender-balanced groups: MBSR (N=90, average age 46.6 ± 11.8 years, 53 female), WL (N=84, average age 46.0 ± 11.7 years, 53 female) or the HEP active control intervention (N=90, average age 45.4 ± 12.5 years, 55 female), which has been validated in a separate study in which the intervention procedures are described in further detail (MacCoon *et al*., 2012). (See Table 1 for detailed demographic information for each group.) All study staff, except the participant coordinator and project manager (and their undergraduate assistants), were blind to group assignment. Blinded group indicators were used, and only the participant coordinator had access to the key.

**Table 1.** Detailed demographic information.

|  |  | Dataset 1 |  |  | Dataset 2 |  |  |
| --- | --- | --- | --- | --- | --- | --- | --- |
|  |  | MBSR<br>(n=35) | HEP<br>(n=36) | WL<br>(n=35) | MBSR<br>(n=40) | HEP<br>(n=37) | WL<br>(n=35) |
| Age | Mean | 50.2 | 47.9 | 48.4 | 44.8 | 44.2 | 45.2 |
|  | SD | 9.4 | 12.5 | 10.5 | 13.3 | 12.8 | 12.5 |
|  | Minimum | 26 | 26 | 26 | 25 | 25 | 25 |
|  | Maximum | 65 | 66 | 65 | 64 | 65 | 65 |
| Sex | Female | 23 | 20 | 23 | 20 | 23 | 23 |
|  | Male | 12 | 16 | 12 | 20 | 14 | 12 |
| Race | American Indian/<br>Alaska Native | 1 | 0 | 0 | 1 | 1 | 1 |
|  | Asian | 1 | 1 | 0 | 5 | 1 | 6 |
|  | Black/ African<br>American | 0 | 0 | 0 | 0 | 2 | 0 |
|  | Native Hawaiian/<br>Pacific Islander | 0 | 0 | 0 | 0 | 0 | 1 |
|  | Multi-racial | 1 | 1 | 1 | 2 | 1 | 3 |
|  | White | 32 | 34 | 32 | 30 | 31 | 24 |
|  | Declined response | 0 | 0 | 2 | 2 | 1 | 0 |
| Ethnicity | Hispanic | 1 | 1 | 6 | 3 | 3 | 1 |
|  | Not Hispanic | 34 | 35 | 29 | 37 | 33 | 34 |

Six participants were excluded due to brain abnormalities, and two participants were missing structural data due to technical difficulties, resulting in 256 participants (average age 45.7 ± 12.0 years, 156 female) with baseline (T1) structural MRI data. Eighteen participants withdrew prior to T2 data collection (8 MBSR, 1 HEP, 9 WL), thirteen participants were excluded because they failed to attend a minimum of 2 classes for the assigned intervention (9 HEP, 4 MBSR), and seven participants were missing T2 structural MRI data due to technical difficulties, resulting in 75 MBSR (average age 47.3 ± 11.9 years, 43 female), 73 HEP (average age 46.0 ± 12.7 years, 43 female), and 70 WL (average age 46.8 ± 11.6 years, 46 female) participants with T2 structural MRI data.

The MBSR courses were delivered by experienced and certified MBSR instructors and consisted of practices and teachings aimed at increasing mindfulness, including yoga, meditation, and body awareness. The HEP course served as an active control, which was matched to MBSR and consisted of exercise, music therapy, and nutrition education and practices. The intervention and randomization procedures were identical to those detailed by MacCoon et al (2012). MBSR and HEP participants recorded logs of the minutes they spent each day on the respective practices at home (i.e., outside of class), which were summed to calculate a variable for total minutes of practice for each participant (except those in the WL group). Classes for both interventions (MBSR and HEP) were taught through the University of Wisconsin Health, Integrative Health department in Madison, Wisconsin.

### Data collection

Participants completed a baseline data collection visit prior to randomization, and a second visit following the 8-week intervention period. Both visits took place at the Waisman Laboratory for Brain Imaging and Behavior at the University of Wisconsin – Madison. The second sample of MNP also completed a third, long-term follow-up session approximately six months after the second visit that was not included in the current analysis. At each visit, participants attended a 24-hour lab session that included an MRI scan and the Five Facet Mindfulness Questionnaire (FFMQ) (Baer *et al*., 2008), among other measures as part of a larger multi-session, multi-project study. We examined the FFMQ to gauge the efficacy of MBSR for improving mindfulness, given its use in the prior studies that we attempt to replicate here (Hölzel *et al*., 2011), and despite its apparent limitations (Baer *et al*., 2008; Davidson and Kaszniak, 2015; Goldberg *et al*., 2015; Van Dam *et al*., 2017). Experimenters were blind to the group assignment during data collection. UW – Madison’s Health Sciences Institutional Review Board approved the protocol, and all participants provided consent and were given monetary compensation for their participation.

### Image Acquisition

Anatomical images for sample one were acquired on a GE X750-3.0 Tesla MRI scanner device with an 8-channel head coil, and consisted of a high-resolution 3D T1-weighted inversion recovery fast gradient echo image (inversion time = 450 msec, 256×256 in-plane resolution, 256 mm FOV, 124×1.0 mm axial slices). Anatomical images for sample two were acquired on the same scanner using a 32-channel head coil with the same scan sequence, except with 192×1.0 mm axial slices.

### Anatomical Image Processing

Image processing was conducted in FreeSurfer using the automated longitudinal pipeline (stable release version 6.0), which included skull-stripping, registration, intensity normalization, Talairach transformation, tissue segmentation, and surface tessellation (Fischl and Dale, 2000; Reuter *et al*., 2012). Hand edits were conducted to correct errors in the automated processing, primarily to the base, and if needed, to the subsequently generated longitudinal images. Manual edits in the base included editing the Talairach registration, wm.mgz, and brainmask.mgz volumes. Edits to the Talairach registration occurred when the initial registration was a poor fit to the subject brain. In both the base and longitudinal phases, control points were added to correct intensity normalization errors and white matter omissions.

Additionally, if the white and pial surfaces did not follow white and gray matter boundaries, voxel edits were made on the wm.mgz and brainmask.mgz volumes, respectively. Images were resampled to fsaverage space using the FreeSurfer program mris_preproc, and difference maps were generated for each subject (T2-T1) using the “paired-diff” option. The resulting difference maps were then smoothed to 8 mm full-width half maximum with mri_surf2surf, and used as inputs for subsequent group analysis.

FreeSurfer’s automated brain segmentation tool (Aseg) (Fischl and Dale, 2000) was used to extract measures of GMV from sub-cortical regions (see Figure 1c for depiction of the right amygdala ROI), and the Desikan-Killany atlas was used to extract GMV for the insula (Desikan *et al*., 2006). A mask of clusters with significant change in MBSR participants was provided by Hölzel (Hölzel *et al*., 2011), which was registered to individual subject space using the FreeSurfer command mri_label2vol using transformation matrices generated for each subject with the FreeSurfer command mni152reg. The resultant TPJ, PCC and 2 cerebellar masks (depicted in the MNI152 template space in Figure 2) were then used to extract GMV measures for each subject using the FreeSurfer program mris_segstats. We masked each of these cortical ROIs to exclude white matter based on each subject’s white matter segmentation, as generated in the FreeSurfer processing pipeline with the program recon-all. The significance of results does not change using the original ROIs, without additional masking.

**Figure 2.**
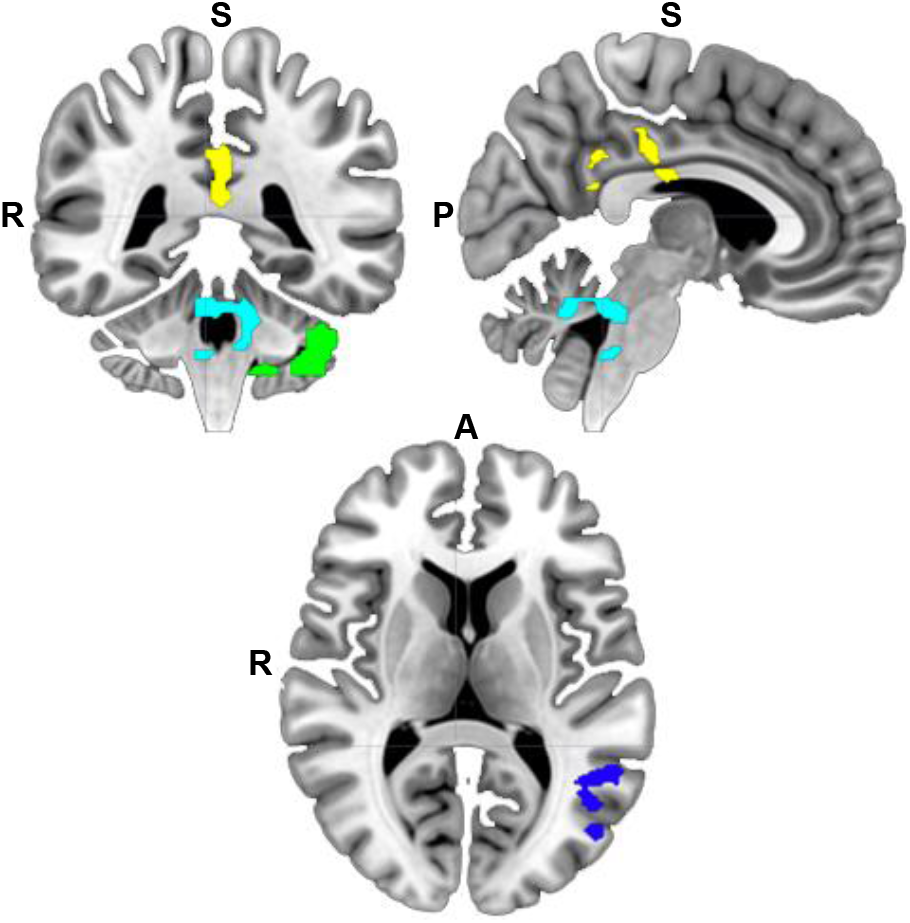
Masks defined from prior research. Regions of interest (ROIs) for posterior cingulate (yellow), left temporoparietal junction (dark blue), cerebellum (green), and cerebellum/ brainstem (teal) were defined based on a thresholded statistical map in which increased gray matter volume was previously reported following Mindfulness-Based Stress Reduction (Hölzel *et al*., 2011). White matter was masked from the cortical ROIs for each subject prior to extraction of GMV. R=right; A=anterior; P=posterior; S=superior

We also completed sensitivity analysis with multiple imputation for ROI analyses, and with data processed using Statistical Parametric Mapping (SPM) software to verify that the absence of significant results and lack of replication of prior work were not due to differences in software or the specific structural measures employed (e.g., GMV versus GMD). Details for multiple imputation and SPM pipelines are available in the Supplementary Information and included both standard SPM12 and SPM12 Computational Anatomy Toolbox (CAT12) longitudinal pipelines, the latter being calibrated to detect smaller changes in brain structure than earlier software. Full results of sensitivity analysis are presented in Supplementary Tables 3-8.

### Statistical Analysis – Voxelwise

Whole-brain analysis was performed on GMV and CT difference maps (T2-T1) using the FreeSurfer program mri_glmfit with a single, 3-level variable of interest to model Group (MBSR, HEP or WL) with covariates to control for age, gender, total (whole-brain) GMV, and sample. The FreeSurfer program mri_glmfit-sim was then used to correct for multiple comparisons with gaussian random fields and an added correction to control for comparisons across the two hemispheres. Analysis of the effect of home practice included an additional regressor with total home practice minutes and modeled a contrast for the interaction of Group x Practice.

### Statistical Analysis – Regions of interest

All ROI analyses were performed using the lm function in R statistics software (R Core Team, 2013), and *p*-value computation used the modelSummary function of the lmSupport package (Curtin, 2015). The GMV difference (T2-T1) for each ROI was regressed (separately) on Group with covariates to control for participant age, gender, sample (i.e., one versus two), and total whole-brain GMV. Analysis of the impact of home practice time on GMV included the addition of total home practice minutes and its interaction with Group. Outliers were identified based on Cook’s D using a cutoff threshold of 4/(N-P), where N and P correspond to the sample size and number of model parameters, respectively, and removed from analyses. The number of outliers per group ranged from two to eight for MBSR, from three to six for HEP, and from one to six for WL. We used a false discovery rate (FDR) correction to control for multiple comparisons across all 12 ROIs using the p.adjust function. Corrected *p*-values are indicated by *p*\* in the text. A summary of descriptive statistics for average GMV for all ROIs is presented in Table 2, and results of all statistical tests are presented in Supplementary Tables 1 to 8.

**Table 2.**
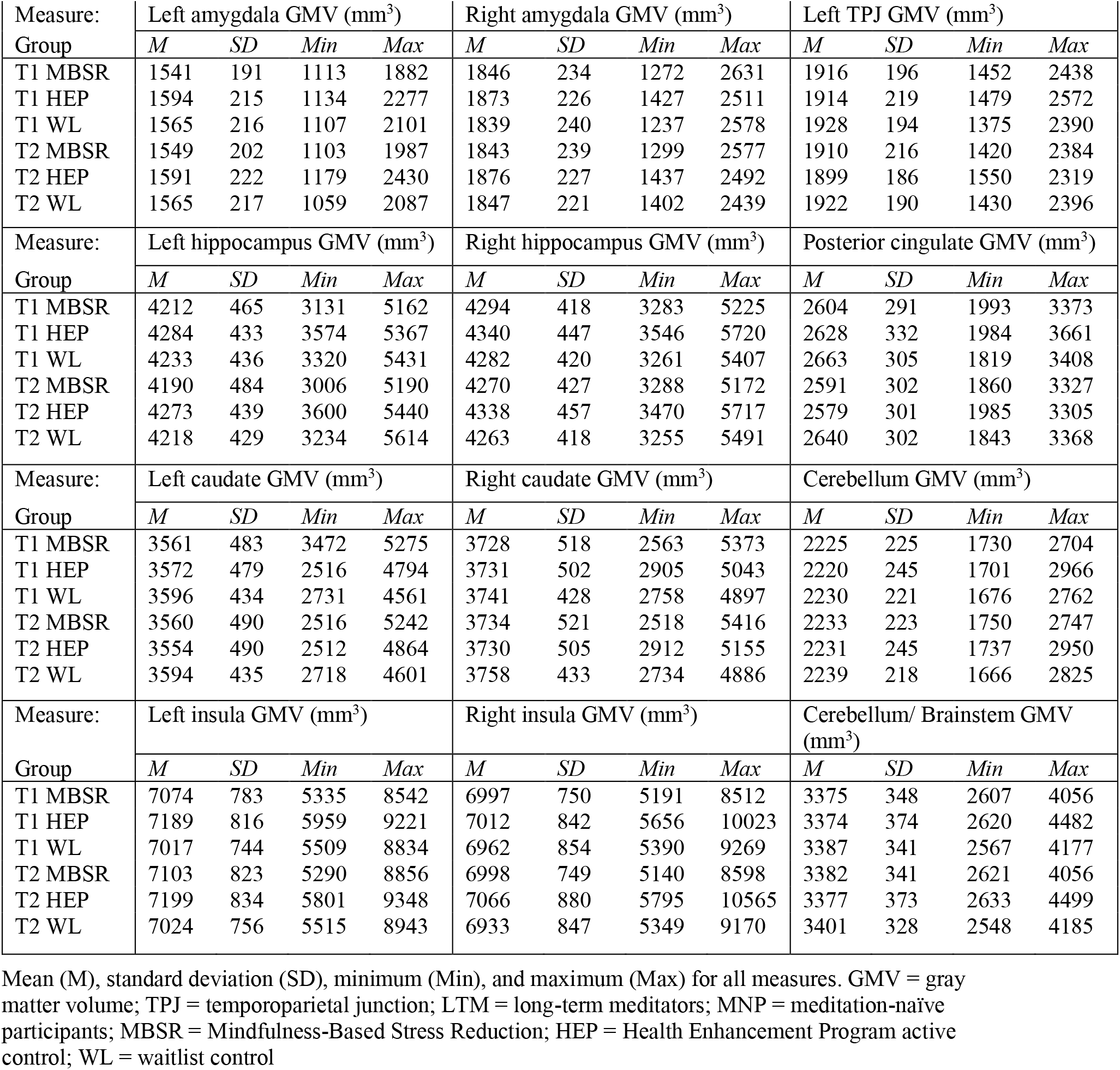
Descriptive statistics.

## Supporting information

CONSORT checklist

## Data Availability

Data are available upon request to the corresponding author. Un-thresholded statistical maps are available on NeuroVault at this link: https://identifiers.org/neurovault.collection:7634

https://osf.io/jrpnq/

## Data Availability

https://osf.io/jrpnq/

## Code Availability

Code for the data processing and analysis can be found at the Open Science Framework at this link: https://osf.io/jrpnq/

## Acknowledgments

This work was supported by the National Center for Complementary and Alternative Medicine (NCCAM) P01AT004952 to RJD, grants from the National Institute of Mental Health (NIMH) R01-MH43454, P50-MH084051 to RJD, grants from the Fetzer Institute and the John Templeton Foundation to RJD, core grants to the Waisman Center from the National Institute of Child Health and Human Development [P30 HD003352-449015, U54 HD090256]. TRAK was supported by the National Institute of Mental Health award number T32MH018931.

We would like to thank Michael Anderle, Ron Fisher, Lisa Angelos, Heather Hessenthaler, Trina Nelson, Amelia Cayo, Michael Kruepke, Jon Brumbaugh, Sarah Christens, Jane Sachs, Jeanne Harris, Mariah Brown, Elizabath Nord, Gina Bednarek, Kara Chung, Pema Lhamo, David Bachhuber, Amelia Cayo, Christopher Harty, Sonam Kindy and Dan Dewitz for assistance with recruitment and/or data collection. We would also like to thank Katherine Bonus, Devin Coogan, Bob Gillespie, Diana Grove, Lori Gustafson, Matthew Hirshberg, Peggy Kalscheur, Chad McGehee, Vincent Minichiello, Laura Pinger, Lisa Thomas Prince, Kristi Rietz, Sara Shatz, Chris Smith, Heather Sorensen, Jude Sullivan, Julie Thurlow, Michael Waupoose, Sandy Wojtal-Weber and Pam Young for teaching and/or coordinating the interventions, and John Koger, Ty Christian, David Thompson, John Ollinger, Martina Ly, Nate Vack, and Andy Schoen for technical assistance.

## Author Contributions

T.R.A.K., K.D., and R.H. contributed to data collection. T.R.A.K., K.D., R.H., C.K., and L.Y.T. processed the data. T.R.A.K. analyzed the data and wrote the manuscript. C.K. and R.G. contributed to planning the analysis. M.H. conducted the multiple imputation analysis. R.G., M.A.R., and A.L. contributed to the design of the study. R.J.D. designed and supervised the study. All authors contributed to writing the manuscript.

