## Supplementary Materials for "Non-replication of structural brain changes from Mindfulness-Based Stress Reduction: Two combined randomized controlled trials"

**This PDF file includes:**

Supplementary Methods
Figures S1 to S2
Tables S1 to S8

### Supplementary Methods

*Multiple Imputation*

For intention-to-treat analyses using all assigned participants, we conducted multivariate imputation through chained equations using the MICE package in R[1–3]. We imputed 50 complete datasets using predictive mean matching or logistic regression methods depending on the variable (i.e., continuous or categorical). All variables used in analyses along with baseline scores on anxiety and negative affect were entered into the imputation model as predictors to improve estimation. Due to high correlations between time 1 and time 2 measures of brain structure, we transformed then imputed change score outcomes (i.e., *JAV*[1]). We confirmed imputation plausibility by plotting the range of imputed values against the range of observed values, the distribution of the imputed values against the distribution of the observed values, and the observed versus imputed values. Convergence was confirmed through plotting the mean and separately the standard deviation of each iteration of imputation for each imputed outcome. We then estimated regression models including all standard covariates for each outcome on each imputed dataset, and pooled results according to Rubin's rules[4]. Full results are in Tables S3-4.

*Statistical Parametric Mapping (SPM) 12 image processing & analysis*

Structural (T1) images were manually realigned to the anterior and posterior commissures (AC-PC), which included adjusting the roll, pitch, and yaw until the AC and PC were in the same axial plane in the sagittal view, and the midlines were oriented vertically in the coronal and axial views. Following manual realignment, T1 images processed according to the longitudinal pipeline in SPM12 (http://www.fil.ion.ucl.ac.uk/spm. Each participant's Time 1 (baseline) and Time 2 (post-intervention period) scans were then registered using pairwise inverse-consistent alignment in SPM12, including bias field correction, and which generates a subject average (spatial mid-point) image and the Jacobian rate (the difference between the Jacobian determinants for each scan when registered to the mid-point average, divided by the time between scans)[5]. The average T1 image for each participant was then segmented into gray matter, white matter, and cerebrospinal fluid. The gray matter segmentation of the average T1 from the prior step was then multiplied by the Jacobian rate to give the rate of change in each participant's average space. A study-specific average space was then generated using the DARTEL (Diffeomorphic Anatomical Registration using Exponentiated Lie Algebra) algorithm[6], and the participants' T1 images from the prior step aligned and normalized to Montreal Neurological Insitute (MNI) space through the group template. Finally, images were modulated to preserve volume and smoothed with a 8 mm full-width at half-maximum (FWHM) Gaussian kernel[7].

Analysis of rate of change in gray matter density (GMD) from SPM12 was conducted using the SPM12 Estimate function. Wholebrain, voxelwise analysis was conducted using a factorial model with a single, 3-level factor for Group, and covariates to control for participant age, gender, sample, and total gray matter volume. Wholebrain statistical maps for each contrast (e.g., MBSR-HEP) were corrected using Family-Wise Error (FWE) and thresholded at $p<0.05$. Average GMD rate of change was extracted from each ROI and regressed on Group in a linear model including the same covariates as whole-brain analysis. Analysis of home practice time built from the models of group differences by adding the interaction with home practice time, and thus comparisons were limited to MBSR and HEP. Full results of ROI analysis are in Tables S5-6.

*SPM-Computational Anatomy Toolbox 12 (CAT12) image processing & analysis*

The manually realigned T1 images that were used in the SPM12 longitudinal processing pipeline (described above), were additionally processed through the CAT12 longitudinal pipeline that was developed with higher sensitivity to detect smaller changes in brain structure, over relatively shorter periods of time (e.g. scans less than one year apart) compared with the standard SPM12 pipeline[8,9]. We used the automated cat_batch_long script, which included modified steps from the standard SPM12 processing pipeline, including tissue segmentation, bias field correction, registration to MNI standard space using the DARTEL algorithm, and modulation to preserve volume. However, while the CAT12 longitudinal pipeline included co-registration of each participants scans, the output from longitudinal processing produced separate files for Time 1 and Time 2, which were entered separately into group analysis. Following the standard CAT12 longitudinal processing, images were smoothed with an 8 mm FWHM Gaussian kernel.

Wholebrain, voxelwise analysis was conducted using a flexible factorial model with 3 factors: Subject, Group, and Time; and the interaction of Group x Time. Participant age, gender, study, and total gray matter volume were included as covariates of no-interest. Contrasts maps were generated for the Group x Time interactions for MBSR versus HEP, and for MBSR versus WL, corrected for multiple comparisons using FWE, and thresholded at $p<0.05$. Average GMD was extracted from each scan (T1 and T2 for each participant, separately) for each ROI using the CAT12 toolbox. ROI analysis was conducted by regressing the GMD differences scores (T2-T1) for each region on Group and including the same covariates as wholebrain analysis. Similar to analysis of SPM12 data, analysis of home practice time built from the models of group differences by adding the interaction with home practice time, and thus comparisons were limited to MBSR and HEP. Full results of ROI analysis are in Table S7-8.

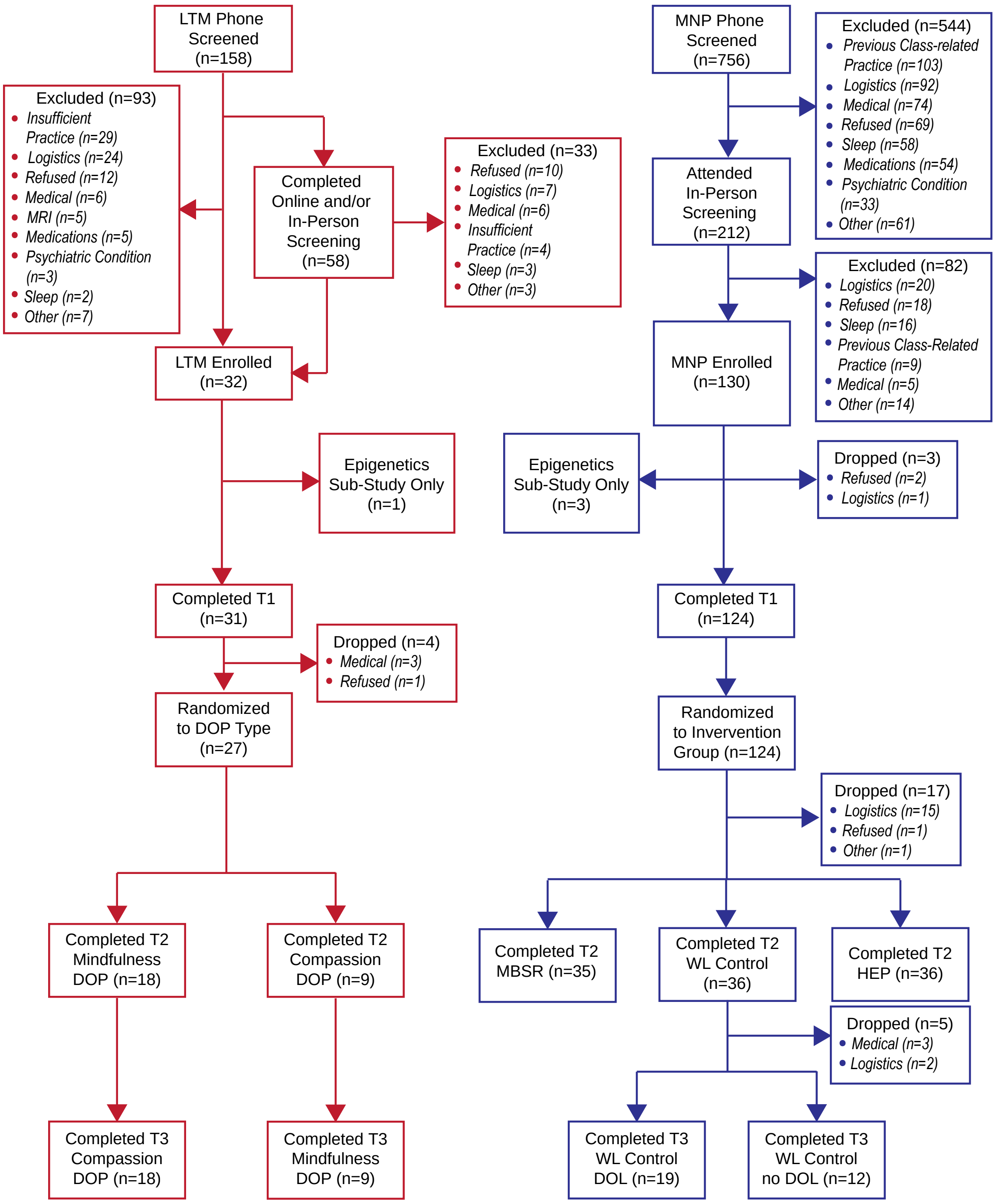


**Fig.**

**S1.** Dataset 1 CONSORT diagram. The current study includes data from the randomized-controlled trial of meditation-naïve participants (MNP), shown in blue. Participants completed a baseline visit (T1) prior to randomization to either Mindfulness-Based Stress Reduction (MBSR), the Health Enhancement Program (HEP) active control intervention, or a waitlist control (WL) group. Participants completed a post-intervention (T2) visit, and the WL group also completed a third visit (T3) as a control for the long-term meditators (LTM) in a separate arm of the trial.

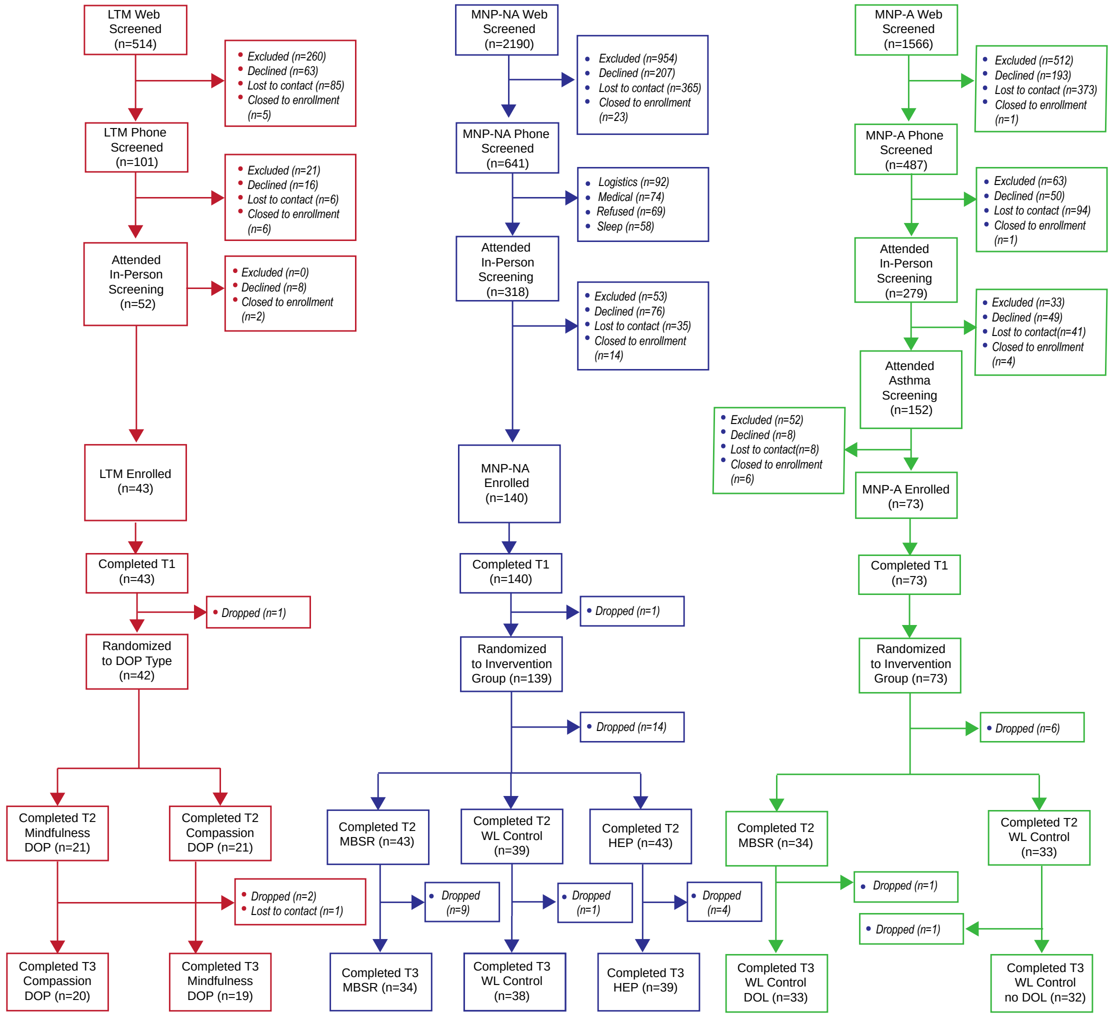


**Fig. S2.** Dataset 2 CONSORT diagram. The current study includes data from the randomized-controlled trial of non-asthmatic, meditation-naïve participants (MNP-NA), shown in blue in the middle. Participants completed a baseline lab visit (T1) prior to randomization to either Mindfulness-Based Stress Reduction (MBSR), the Health Enhancement Program (HEP) active control intervention, or a waitlist control (WL) group. Participants completed a post-intervention (T2) lab visit, and a third lab visit for long-term follow-up (T3). Long-term meditators (LTM) and meditation-naïve asthmatics (MNP-A) were not included in the current study, which focused on replicating prior studies on the impact of MBSR.

**Table S1.** Statistics for analysis of change in regional gray matter volume.

| Brain region GMV ($mm^3$) | Comparison (T2-T1) | Statistic | | | | | |
|---|---|---|---|---|---|---|---|
| | | *df* | *t* | *CI* | $p\eta^2$ | *p* | *p** |
| Left amygdala | MBSR vs HEP | 198 | -1.65 | [-31.79:2.84] | 0.02 | 0.10 | 0.53 |
| | MBSR vs WL | 198 | -1.40 | [-29.54:5.03] | 0.02 | 0.16 | 0.57 |
| Right amygdala | MBSR vs HEP | 198 | -0.13 | [-19.17:16.88] | <0.01 | 0.90 | 0.96 |
| | MBSR vs WL | 198 | 0.38 | [-14.83:21.8] | <0.01 | 0.71 | 0.77 |
| Left insula | MBSR vs HEP | 200 | 0.05 | [-39.79:41.84] | <0.01 | 0.96 | 0.96 |
| | MBSR vs WL | 200 | -0.71 | [-56.56:26.72] | <0.01 | 0.48 | 0.65 |
| Right insula | MBSR vs HEP | 198 | 0.26 | [-40.03:52.40] | 0.01 | 0.79 | 0.96 |
| | MBSR vs WL | 198 | -1.19 | [-73.95:18.39] | 0.01 | 0.24 | 0.58 |
| Left caudate | MBSR vs HEP | 198 | -1.37 | [-38.96:7.00] | 0.01 | 0.17 | 0.53 |
| | MBSR vs WL | 198 | -0.94 | [-34.79:12.32] | 0.01 | 0.35 | 0.65 |
| Right caudate | MBSR vs HEP | 207 | 0.17 | [-19.24:22.84] | 0.02 | 0.87 | 0.96 |
| | MBSR vs WL | 207 | 1.75 | [-2.33:39.91] | 0.02 | 0.08 | 0.54 |
| Left hippocampus | MBSR vs HEP | 201 | 0.94 | [-13.4:37.87] | <0.01 | 0.35 | 0.70 |
| | MBSR vs WL | 201 | 0.69 | [-16.72:34.76] | <0.01 | 0.49 | 0.65 |
| Right hippocampus | MBSR vs HEP | 199 | 2.06 | [1.01:47.66] | 0.03 | 0.04 | 0.48 |
| | MBSR vs WL | 199 | -0.01 | [-23.49:23.18] | 0.03 | 0.99 | 0.99 |
| Left TPJ | MBSR vs HEP | 196 | 1.32 | [-12.46:62.83] | 0.01 | 0.19 | 0.53 |
| | MBSR vs WL | 196 | 0.70 | [-23.94:50.56] | 0.01 | 0.48 | 0.65 |
| Posterior Cingulate | MBSR vs HEP | 198 | -1.23 | [-103.92:24] | 0.01 | 0.22 | 0.53 |
| | MBSR vs WL | 198 | -1.30 | [-104.24:21.27] | 0.01 | 0.19 | 0.57 |
| Cerebellum | MBSR vs HEP | 199 | -0.47 | [-13.58:8.31] | <0.01 | 0.64 | 0.96 |
| | MBSR vs WL | 199 | 0.43 | [-8.53:13.33] | <0.01 | 0.67 | 0.77 |
| Cerebellum/ Brainstem | MBSR vs HEP | 204 | 0.23 | [-14.63:18.47] | 0.02 | 0.82 | 0.96 |
| | MBSR vs WL | 204 | 1.72 | [-2.12:31.13] | 0.02 | 0.09 | 0.54 |

T2 = post-intervention measure; T1 = baseline/pre-intervention measure; MBSR = Mindfulness-Based Stress Reduction; HEP = Health Enhancement Program active control; WL = waitlist control; GMV = gray matter volume; *df* = degrees of freedom; *CI* = confidence interval; TPJ = temporoparietal junction

**Table S2.** Statistics for analysis of MBSR versus HEP practice time and change in GMV (T2-T1).

| Brain region (GMV [mm$^3$]; T2-T1) X Practice (min.) interaction | Statistic | | | | | |
|---|---|---|---|---|---|---|
| | *df* | *t* | *CI* | p$\eta^2$ | *p* | *p** |
| Left amygdala: MBSR vs HEP | 130 | -1.73 | [-0.03:0.00] | 0.02 | 0.09 | 0.26 |
| **Right amygdala: MBSR v HEP** | **128** | **-3.71** | **[-0.05:-0.01]** | **0.10** | **<0.01** | **<0.01** |
| MBSR | 64 | -1.59 | [-0.05:-0.01] | 0.04 | 0.12 | 0.63 |
| HEP | 63 | 0.58 | [-0.01:0.01] | 0.01 | 0.57 | 0.74 |
| Left insula | 131 | 1.82 | [0.00:0.06] | 0.02 | 0.07 | 0.26 |
| Right insula | 127 | 0.66 | [-0.03:0.06] | <0.01 | 0.51 | 0.68 |
| Left caudate | 132 | -0.80 | [-0.03:0.01] | <0.01 | 0.43 | 0.68 |
| Right caudate | 136 | -0.02 | [-0.02:0.01] | <0.01 | 0.98 | 0.98 |
| Left hippocampus | 132 | 1.58 | [0.00:0.03] | 0.02 | 0.12 | 0.28 |
| Right hippocampus | 131 | 0.67 | [-0.01:0.02] | <0.01 | 0.50 | 0.68 |
| Left TPJ | 127 | 0.66 | [-0.03:0.05] | <0.01 | 0.51 | 0.68 |
| Posterior Cingulate | 127 | 2.13 | [0.01:0.14] | 0.03 | 0.04 | 0.21 |
| Cerebellum | 131 | -0.39 | [-0.01:0.01] | <0.01 | 0.70 | 0.83 |
| Brainstem | 132 | 0.18 | [-0.01:0.01] | <0.01 | 0.85 | 0.93 |

MBSR = Mindfulness-Based Stress Reduction; HEP = Health Enhancement Program active control; GMV = gray matter volume; T2 = post-intervention measure; T1 = baseline/pre-intervention measure; TPJ = temporo-parietal junction; *df* = degrees of freedom; *CI* = confidence interval

**Table S3.** Statistics for analysis of change of regional gray matter volume using multiple imputation.

| Brain region GMV ($mm^3$) | Comparison (T2-T1) | Statistic | | | | | |
|---|---|---|---|---|---|---|---|
| | | *df* | *t* | *CI* | $p\eta^2$ | *p* | *p** |
| Left amygdala | MBSR vs HEP | 192.21 | -1.11 | [-32.54, 9.04] | 0.01 | 0.27 | 0.98 |
| | MBSR vs WL | 207.38 | -0.74 | [-28.64, 13.03] | <0.01 | 0.46 | 0.98 |
| Right amygdala | MBSR vs HEP | 196.22 | 0.83 | [-12.80, 31.52] | <0.01 | 0.41 | 0.98 |
| | MBSR vs WL | 201.25 | 1.07 | [-19.34, 34.71] | 0.01 | 0.29 | 0.98 |
| Left insula | MBSR vs HEP | 172.26 | -0.32 | [-59.35, 42.98] | <0.01 | 0.75 | 0.98 |
| | MBSR vs WL | 192.53 | -0.95 | [-75.24, 26.48] | <0.01 | 0.35 | 0.98 |
| Right insula | MBSR vs HEP | 192.00 | 1.75 | [-6.63, 112.34] | 0.02 | 0.08 | 0.97 |
| | MBSR vs WL | 177.09 | -1.12 | [-97.35, 27.06] | 0.01 | 0.27 | 0.98 |
| Left caudate | MBSR vs HEP | 184.78 | -0.96 | [-43.28, 14.94] | 0.01 | 0.34 | 0.98 |
| | MBSR vs WL | 196.97 | -0.03 | [-29.70, 28.94] | <0.01 | 0.98 | 0.98 |
| Right caudate | MBSR vs HEP | 215.08 | -0.33 | [-31.62, 22.50] | <0.01 | 0.74 | 0.98 |
| | MBSR vs WL | 204.89 | 0.65 | [-18.87, 22.50] | <0.01 | 0.52 | 0.98 |
| Left hippocampus | MBSR vs HEP | 207.78 | 0.92 | [-15.14, 41.86] | <0.01 | 0.36 | 0.98 |
| | MBSR vs WL | 192.66 | 0.68 | [-19.57, 40.01] | <0.01 | 0.50 | 0.98 |
| Right hippocampus | MBSR vs HEP | 201.30 | 1.77 | [-2.69, 49.72] | 0.02 | 0.08 | 0.97 |
| | MBSR vs WL | 204.06 | 0.42 | [-21.10, 32.35] | <0.01 | 0.68 | 0.98 |
| Left TPJ | MBSR vs HEP | 205.78 | 0.25 | [-48.18, 62.00] | <0.01 | 0.81 | 0.98 |
| | MBSR vs WL | 210.29 | -0.13 | [-59.84, 52.25] | <0.01 | 0.89 | 0.98 |
| Posterior Cingulate | MBSR vs HEP | 204.43 | -0.05 | [-91.01, 86.31] | <0.01 | 0.96 | 0.98 |
| | MBSR vs WL | 208.16 | -0.43 | [-110.08, 70.51] | <0.01 | 0.67 | 0.98 |
| Cerebellum | MBSR vs HEP | 203.67 | -0.94 | [-21.54, 7.66] | <0.01 | 0.92 | 0.98 |
| | MBSR vs WL | 207.93 | -0.64 | [-19.65, 10.07] | <0.01 | 0.889 | 0.98 |
| Cerebellum/ Brainstem | MBSR vs HEP | 220.10 | -0.94 | [-27.35, 9.65] | 0.01 | 0.63 | 0.98 |
| | MBSR vs WL | 230.75 | 0.15 | [-17.20, 20.09] | <0.01 | 0.62 | 0.98 |

T2 = post-intervention measure; T1 = baseline/pre-intervention measure; MBSR = Mindfulness-Based Stress Reduction; HEP = Health Enhancement Program active control; WL = waitlist control; GMV = gray matter volume; *df* = degrees of freedom; *CI* = confidence interval; TPJ = temporoparietal junction

**Table S4.** Statistics for MI analysis of MBSR versus HEP practice time and change in GMV.

| Brain region (GMV [mm$^3$]; T2-T1) X Practice (min.) interaction | Statistic | | | | | |
|---|---|---|---|---|---|---|
| | *df* | *t* | *CI* | p$\eta^2$ | *p* | *p** |
| Left amygdala | 143.03 | 0.71 | [-0.01, 0.02] | 0.01 | 0.478 | .637 |
| Right amygdala | 143.03 | 1.89 | [-0.001, 0.03] | 0.02 | 0.061 | .340 |
| Left insula GMV | 143.03 | -1.25 | [-0.06, 0.01] | 0.02 | .214 | .514 |
| Right insula | 143.03 | -1.85 | [-0.08, .003] | 0.02 | .066 | .340 |
| Left caudate | 143.03 | 0.20 | [-0.02, 0.02] | <0.01 | .844 | .921 |
| Right caudate | 143.03 | -1.55 | [-0.04, 0.005] | 0.02 | .123 | .369 |
| Left hippocampus | 143.03 | -0.85 | [-0.03, 0.01] | 0.01 | .397 | .560 |
| Right hippocampus | 143.03 | -0.58 | [-0.02, 0.01] | 0.01 | .566 | .679 |
| Left TPJ | 143.03 | 0.92 | [-0.02, 0.06] | 0.01 | .359 | .560 |
| Posterior Cingulate | 143.03 | 0.04 | [-0.11, 0.03] | <0.01 | .315 | .560 |
| Cerebellum | 143.03 | 1.73 | [-0.001. 0.02] | 0.03 | .085 | .340 |
| Brainstem | 143.03 | -0.004 | [-0.01. 0.01] | <0.01 | .997 | .997 |

MI = multiple imputation; T2 = post-intervention measure; T1 = baseline/pre-intervention measure; MBSR = Mindfulness-Based Stress Reduction; HEP = Health Enhancement Program active control; GMV = gray matter volume; TPJ = temporo-parietal junction; *df* = degrees of freedom; *CI* = confidence interval

**Table S5.** Statistics for analysis of rate of change of regional gray matter density from SPM12.

| Brain region (GMD) | Comparison (T2-T1) | Statistic | | | | | |
|---|---|---|---|---|---|---|---|
| | | *df* | *t* | *CI* | p$\eta^2$ | *p* | *p** |
| Left amygdala | MBSR vs HEP | 198 | -0.74 | [-5.73:2.59] | <0.01 | 0.46 | 0.61 |
| | MBSR vs WL | 198 | -0.44 | [-5.14:3.25] | <0.01 | 0.66 | 0.94 |
| Right amygdala | MBSR vs HEP | 201 | -0.24 | [-4.91:3.86] | <0.01 | 0.81 | 0.83 |
| | MBSR vs WL | 201 | -0.18 | [-4.73:3.94] | <0.01 | 0.86 | 0.94 |
| Left insula | MBSR vs HEP | 197 | -0.57 | [-29.12:16.09] | 0.01 | 0.57 | 0.68 |
| | MBSR vs WL | 197 | 1.11 | [-9.86:35.14] | 0.01 | 0.27 | 0.94 |
| Right insula | MBSR vs HEP | 197 | -1.97 | [-40.53:0.06] | 0.02 | 0.05 | 0.45 |
| | MBSR vs WL | 197 | -1.04 | [-31.68:9.76] | 0.02 | 0.30 | 0.94 |
| Left caudate | MBSR vs HEP | 201 | -1.58 | [-18.18:2.02] | 0.01 | 0.12 | 0.45 |
| | MBSR vs WL | 201 | -0.28 | [-11.75:8.80] | 0.01 | 0.78 | 0.94 |
| Right caudate | MBSR vs HEP | 197 | 0.21 | [-9.15:11.32] | <0.01 | 0.83 | 0.83 |
| | MBSR vs WL | 197 | 0.88 | [-5.68:14.93] | <0.01 | 0.38 | 0.94 |
| Left hippocampus | MBSR vs HEP | 200 | -1.18 | [-16.82:4.22] | 0.01 | 0.24 | 0.55 |
| | MBSR vs WL | 200 | -0.05 | [-10.93:10.43] | 0.01 | 0.96 | 0.96 |
| Right hippocampus | MBSR vs HEP | 200 | 0.88 | [-5.91:15.45] | <0.01 | 0.38 | 0.57 |
| | MBSR vs WL | 200 | 0.31 | [-9.11:12.50] | <0.01 | 0.76 | 0.94 |
| Left TPJ | MBSR vs HEP | 199 | -1.46 | [-8.08:1.20] | 0.02 | 0.15 | 0.45 |
| | MBSR vs WL | 199 | 0.49 | [-3.62:5.99] | 0.02 | 0.63 | 0.94 |
| Posterior Cingulate | MBSR vs HEP | 200 | -1.00 | [-5.46:1.79] | 0.02 | 0.32 | 0.55 |
| | MBSR vs WL | 200 | 0.98 | [-1.86:5.55] | 0.02 | 0.33 | 0.94 |
| Cerebellum | MBSR vs HEP | 197 | -1.54 | [-6.23:0.76] | 0.01 | 0.12 | 0.45 |
| | MBSR vs WL | 197 | -0.64 | [-4.67:2.39] | 0.01 | 0.52 | 0.94 |
| Cerebellum/ Brainstem | MBSR vs HEP | 195 | -1.09 | [-7.09:2.05] | 0.01 | 0.28 | 0.55 |
| | MBSR vs WL | 195 | 0.38 | [-3.76:5.53] | 0.01 | 0.71 | 0.94 |

SPM = Statistical Parametric Mapping; MBSR = Mindfulness-Based Stress Reduction; HEP = Health Enhancement Program active control; WL = waitlist control; GMD = gray matter density; T2 = post-intervention measure; T1 = baseline/pre-intervention measure; *df* = degrees of freedom; *CI* = confidence interval; TPJ = temporoparietal junction

**Table S6.** Statistics for SPM12 analysis of MBSR versus HEP practice time and rate of change in GMD.

| Brain region (GMD; T2-T1) X Practice (min.) interaction | Statistic | | | | | |
|---|---|---|---|---|---|---|
| | *df* | *t* | *CI* | p$\eta^2$ | *p* | *p** |
| Left amygdala | 129 | 1.12 | [0.00:0.01] | 0.01 | 0.26 | 0.81 |
| Right amygdala | 133 | -0.21 | [0.00:0.00] | <0.01 | 0.83 | 0.83 |
| Left insula GMV | 130 | 0.60 | [-0.01:0.02] | <0.01 | 0.55 | 0.83 |
| Right insula | 132 | 0.95 | [-0.01:0.02] | 0.01 | 0.34 | 0.81 |
| Left caudate | 134 | -1.01 | [-0.01:0.00] | 0.01 | 0.31 | 0.81 |
| Right caudate | 127 | 1.56 | [0.00:0.02] | 0.02 | 0.12 | 0.73 |
| Left hippocampus | 133 | 2.20 | [0.00:0.02] | 0.04 | 0.03 | 0.36 |
| Right hippocampus | 133 | 0.49 | [-0.01:0.01] | <0.01 | 0.63 | 0.83 |
| Left TPJ | 132 | 0.25 | [0.00:0.00] | <0.01 | 0.81 | 0.83 |
| Posterior Cingulate | 133 | -0.40 | [0.00:0.00] | <0.01 | 0.69 | 0.83 |
| Cerebellum | 129 | 0.37 | [0.00:0.00] | <0.01 | 0.71 | 0.83 |
| Brainstem | 129 | 0.84 | [0.00:0.01] | 0.01 | 0.40 | 0.81 |

SPM = Statistical Parametric Mapping; GMD = gray matter density; MBSR = Mindfulness-Based Stress Reduction; HEP = Health Enhancement Program active control; T2 = post-intervention measure; T1 = baseline/pre-intervention measure; TPJ = temporo-parietal junction; *df* = degrees of freedom; *CI* = confidence interval

**Table S7.** Statistics for analysis of change of regional gray matter density from SPM-CAT12.

| Brain region GM | Comparison (T2-T1) | Statistic | | | | | |
|---|---|---|---|---|---|---|---|
| | | *df* | *t* | *CI* | p$\eta^2$ | *p* | *p** |
| Left amygdala | MBSR vs HEP<br>MBSR vs WL | 198<br>198 | 0.62<br>0.74 | [-6.52:12.49]<br>[-6.08:13.33] | <0.01<br><0.01 | 0.54<br>0.46 | 0.77<br>0.61 |
| Right amygdala | MBSR vs HEP<br>MBSR vs WL | 198<br>198 | 0.19<br>0.83 | [-11.77:14.31]<br>[-7.73:18.90] | <0.01<br><0.01 | 0.85<br>0.41 | 0.85<br>0.61 |
| Left insula | MBSR vs HEP<br>MBSR vs WL | 200<br>200 | 1.74<br>0.88 | [-5.20:82.08]<br>[-24.21:63.60] | 0.01<br>0.01 | 0.08<br>0.38 | 0.48<br>0.61 |
| Right insula | MBSR vs HEP<br>MBSR vs WL | 203<br>203 | 0.60<br>0.56 | [-28.85:53.98]<br>[-30.30:54.14] | <0.01<br><0.01 | 0.55<br>0.58 | 0.77<br>0.70 |
| Left caudate | MBSR vs HEP<br>MBSR vs WL | 199<br>199 | 0.38<br>0.89 | [-14.29:21.10]<br>[-9.97:26.33] | <0.01<br><0.01 | 0.70<br>0.37 | 0.77<br>0.61 |
| Right caudate | MBSR vs HEP<br>MBSR vs WL | 197<br>197 | -0.93<br>0.80 | [-28.12:10.10]<br>[-11.41:26.97] | 0.02<br>0.02 | 0.35<br>0.42 | 0.77<br>0.61 |
| Left hippocampus | MBSR vs HEP<br>MBSR vs WL | 203<br>203 | -0.79<br>0.37 | [-23.67:10.14]<br>[-14.00:20.52] | 0.01<br>0.01 | 0.43<br>0.71 | 0.77<br>0.77 |
| Right hippocampus | MBSR vs HEP<br>MBSR vs WL | 201<br>201 | 0.38<br>0.77 | [-12.07:17.81]<br>[-9.16:21.00] | <0.01<br><0.01 | 0.71<br>0.44 | 0.77<br>0.61 |
| Left TPJ | MBSR vs HEP<br>MBSR vs WL | 198<br>198 | 1.24<br>0.04 | [-1.87:8.16]<br>[-5.03:5.24] | 0.01<br>0.01 | 0.22<br>0.97 | 0.77<br>0.97 |
| Posterior Cingulate | MBSR vs HEP<br>MBSR vs WL | 198<br>198 | -0.55<br>-1.73 | [-26.2:14.79]<br>[-38.91:2.56] | 0.02<br>0.02 | 0.58<br>0.09 | 0.77<br>0.61 |
| Cerebellum | MBSR vs HEP<br>MBSR vs WL | 201<br>201 | 0.47<br>-1.39 | [-7.50:12.25]<br>[-17.17:2.97] | 0.02<br>0.02 | 0.64<br>0.17 | 0.77<br>0.61 |
| Cerebellum/<br>Brainstem | MBSR vs HEP<br>MBSR vs WL | 197<br>197 | -1.74<br>-1.14 | [-13.34:0.85]<br>[-11.51:3.05] | 0.02<br>0.02 | 0.08<br>0.25 | 0.48<br>0.61 |

SPM-CAT = Statistical Parametric Mapping Computational Anatomy Toolbox; MBSR = Mindfulness-Based Stress Reduction; HEP = Health Enhancement Program active control; WL = waitlist control; GMD = gray matter density; T2 = post-intervention measure; T1 = baseline/pre-intervention measure; *df* = degrees of freedom; *CI* = confidence interval; TPJ = temporoparietal junction

**Table S8.** Statistics for SPM-CAT12 analysis of MBSR versus HEP practice time and change in GMD.

| Brain region (GMD; T2-T1) X Practice (min.) interaction | Statistic | | | | | |
|---|---|---|---|---|---|---|
| | *df* | *t* | *CI* | p$\eta^2$ | *p* | *p** |
| Left amygdala | 129 | 0.29 | [-0.01:0.01] | <0.01 | 0.77 | 0.98 |
| **Right amygdala** | **129** | **-2.67** | **[-0.03:0.00]** | **0.05** | **0.01** | **0.10** |
| Left insula GMV | 129 | 0.95 | [-0.02:0.05] | 0.01 | 0.35 | 0.87 |
| Right insula | 132 | -0.09 | [-0.04:0.03] | <0.01 | 0.93 | 0.98 |
| Left caudate | 130 | -0.14 | [-0.01:0.01] | <0.01 | 0.89 | 0.98 |
| Right caudate | 132 | -0.90 | [-0.02:0.01] | 0.01 | 0.37 | 0.87 |
| Left hippocampus | 132 | -0.02 | [-0.01:0.01] | <0.01 | 0.98 | 0.98 |
| Right hippocampus | 129 | -2.03 | [-0.02:0.00] | 0.03 | 0.04 | 0.27 |
| Left TPJ | 129 | -0.71 | [-0.01:0.00] | <0.01 | 0.48 | 0.87 |
| Posterior Cingulate | 132 | 0.67 | [-0.01:0.02] | <0.01 | 0.51 | 0.87 |
| Cerebellum | 133 | -0.37 | [-0.01:0.01] | <0.01 | 0.71 | 0.98 |
| Brainstem | 132 | -0.80 | [-0.01:0.00] | <0.01 | 0.42 | 0.87 |

SPM-CAT = Statistical Parametric Mapping Computational Anatomy Toolbox; MBSR = Mindfulness-Based Stress Reduction; HEP = Health Enhancement Program active control; GMD = gray matter density; TPJ = temporo-parietal junction; *df* = degrees of freedom; *CI* = confidence interval
